# A Mathematical Model of Platelet Antiparasitic Action in Erythrocytes and Merozoites During Malaria Infection

**DOI:** 10.1101/2021.01.25.21250444

**Authors:** Felipe Alves Rubio, Hyun Mo Yang

## Abstract

Platelets have been seen traditionally as fragments of blood mediating coagulation. However, evidence during malaria infection suggests that platelets also act against merozoites, an infectious form of malaria in the bloodstream, and megakaryocytes can release giant platelets with a larger volume than normal platelets. We propose a mathematical model to study the interaction between red blood cells, merozoites, and platelets during malaria infection. We analyzed two cases of the interaction of platelets with malaria infection. In the first one, we considered the isolated action of normal platelets and, in the second one, the joint antiparasitic action of both normal and giant platelets. Numerical simulations were performed to evaluate the stability of the equilibrium points of the system of equations. The model showed that the isolated antiparasitic action of normal platelets corroborates malaria infection control. However, the system can converge to a presence-merozoite equilibrium point, or an oscillatory behavior may appear. The joint antiparasitic action of both normal and giant platelets eliminated the oscillatory behavior and drove the dynamics to converge to lower parasitic concentration than the case of isolated action of normal platelets. Moreover, the joint antiparasitic action of platelets proved more easily capable of eliminating the infection.

## 1 Introduction

In tropical and subtropical regions, diseases transmitted by mosquitoes represent a major public health problem, and malaria is one of them. In 2019, there were 409 thousand deaths by malaria reported by the World Health Organization (WHO). Malaria is caused by protozoan belonging to the genus Plasmodium, which is transmitted by the female mosquito *Anopheles*, being among the diseases that kill the most in the world [1].

During *Anopheles* bite, sporozoites are introduced into the human bloodstream and captured by hepatocytes. After a time in the liver, the sporozoites differentiate into merozoites, which is the parasite form that penetrates erythrocytes and starts the pathogenic phase of malaria infection [5,9]. Inside this infected erythrocyte, after a replication time, more merozoites are released to the bloodstream, and they can infect other naive erythrocytes. This cycle of erythrocyte infection repeats at regular intervals, which is different for each species. The periodic release of merozoites causes typical symptoms like chills, sweating, and fever [6].

Platelets have been traditionally seen as fragments of blood mediating coagulation. However, evidence suggests that platelets are critical in immunology, particularly innate immunity [2,16,19]. They preferentially bind to the infected erythrocytes, and this junction is associated with the death of the parasite [12]. This binding involves interactions between the platelets CD36 molecules and specifically expressed molecules in infected erythrocytes [11]. After binding to infected erythrocytes by merozoites, the platelets are specifically activated and release *α*-granules. The *α*-granule protein, mainly responsible for platelets’ cytocidal action, is the platelet factor 4, denoted by PF4 [13].

Mc Morran et al. (2012) found that both PF4 and Duffy antigen (expressed in erythrocytes) are required to kill the parasite by the action of platelets [13]. Recent studies have shown that PF4 is specifically internalized by the parasite and kills inside the cell. The Duffy antigen is equally essential, and it was discovered that in erythrocytes that lack Duffy antigen, the parasites were not affected by platelets [11].

Lacerda (2010) suggested that during malaria infection, the megakaryocytes can release giant platelets into the bloodstream to compensate for the low absolute number of peripheral platelets. These giant platelets appear to have a shorter time of maturation in detriment of the normal platelets [8]. Thus, platelets, directly on the merozoites or indirectly due to the platelets’ adhesion in the infected cells, may help to control the infection of erythrocyte by merozoites. Here we propose a mathematical model composed of ordinary differential equations to investigate the interaction between platelets and the course of malaria infection. We emphasize that other control mechanisms will not be considered; only the investigation of platelets will be done as an auxiliary response in the controlling of erythrocyte merozoite infection.

The paper is structured as follows. In Section 2, a mathematical model is presented to describe platelets’ action on malaria infection. In Section 3, the model is analyzed in two parts: (i) the model without antiparasitic action of normal and giant platelets, (ii) the effect of antiparasitic action of both normal and giant platelets against the parasite. Section 4 presents a discussion. The conclusion is given in Section 5.

## 2 Model formulation

We assume that uninfected red blood cells, whose concentration is denoted by *H*, are maintained at a constant level (homeostasis) by a constant production rate *K*_1_ and death *µ*_*H*_. These cells, interacting with free merozoites, which are denoted by *X*, are infected at a constant infection rate *β*.

The infected red blood cells, denoted by *H*_*I*_, have an additional mortality rate *µ*_*I*_ due to infection. Each infected red blood cell releases *α* new merozoites in the bloodstream during its average lifetime. The merozoites have a death rate *µ*_*X*_, and *δ*_1_ represents the number of released merozoites that invade uninfected red blood cells.

To describe platelets’ production, we assume a compartment of megakaryocyte precursor cells, denoted by *Z*. We admitted that megakaryocyte precursor cells are produced at a constant rate *K*_2_ and have a death rate *µ*_*Z*_. After a 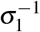 time, they differentiate into megakaryocytes, which concentration is denoted by *M*_1_. This megakaryocyte have death rate *µ*_1_ and, after a *ε*_1_^−1^ time, releases *δ*_2_ normal platelets, which concentration is denoted by *P*_1_, and death rate 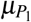.

We consider that, in the presence of infected red blood cells in the bloodstream, with a rate *σ*_2_, the megakaryocyte precursor cell can also be differentiated into a different type of megakaryocyte, denoted by *M*_2_, and produce giant platelets, denoted by *P*_2_. After 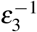 time (less than 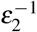), the megakaryocyte *M*_2_ releases *δ*_3_ giant platelets with a larger volume compared to normal platelets. These giant platelets appear during malaria infection to assist the normal platelets, and they have a death rate 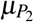.

We also assume that some megakaryocytes can be recruited to produce giant platelets during malaria infection. In this way, we consider that *ε*_2_ represents the recruitment rate of megakaryocyte *M*_1_ to produce giant platelets.

We consider that both normal and giant platelets can act against infected red blood cells to describe platelets’ immune action. The direct action of normal platelets (giant platelets) in merozoites and infected red blood cells is given by the per-capita rate of antiparasitic action *γ*_*X*_ 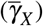 and *γ*_*a*_ 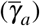, respectively. The parameter *δ*_4_ (*δ*_5_) represents the number of ordinary platelets (giant platelets) bind to the infected red blood cell, and *δ*_6_ (*δ*_7_) represents the number of common platelets (giant platelets) bind to the merozoite.

Based on the above assumptions, we have the following system of differential equations:

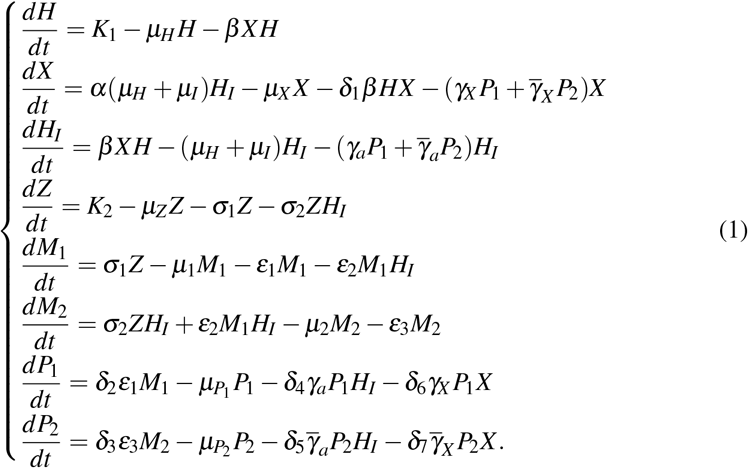

Table 1 summarizes the variables, and Table 2 summarizes the parameters assumed in the model described by the system of equation (1).

**Table 1:** Summary of the model variables (*vol* stands for arbitrary unit of volume, [*H*], [*X*], [*M*] stand for number of, respectively, uninfected or infected red blood cells, merozoites, megakaryocyte precursor cells, and megakaryocytes to produce normal or giant platelets. [*P*] stands for both concentration of normal and giant platelets).

| Symbol | Meaning | Unit |
| --- | --- | --- |
| $H$ | Uninfected red blood cells | $[H]/vol$ |
| $X$ | Merozoites | $[X]/vol$ |
| $H_I$ | Infected red blood cells | $[H]/vol$ |
| $Z$ | Megakaryocyte precursor cells | $[M]/vol$ |
| $M_1$ | Megakaryocytes | $[M]/vol$ |
| $M_2$ | Megakaryocytes producing giant platelets | $[M]/vol$ |
| $P_1$ | Normal platelets | $[P]/vol$ |
| $P_2$ | Giant platelets | $[P]/vol$ |

**Table 2:** Summary of the model parameters (*vol* stands for arbitrary of volume, *T* stands for arbitrary unit of time, [*H*], [*X*], [*Z*], [*M*] stand for number of, respectively, susceptible/infected red blood cells, merozoites, megakaryocyte precursor cells, and megakaryocytes producing normal or giant platelets. [*P*] stands for both concentration of normal and giant platelets). Here we use *vol* = *mm*^3^ and *T* = *day*. ^∗^Values allowed to vary.

| Symbol | Meaning | Value | Unit | Reference |
| --- | --- | --- | --- | --- |
| $K_1$ | Production rate of red blood cells | $6.0 \times 10^4$ | $[H]T^{-1}$ | [3] |
| $\mu_H$ | Mortality rate of red blood cells | $1/120$ | $T^{-1}$ | [3] |
| $\beta$ | Rate of infection red blood cells by merozoite | $*6.0 \times 10^{-8}$ | $[X]^{-1}T^{-1}$ | Assumed |
| $\alpha$ | Number of merozoites released by infected red blood cell | 8 | $[X][H]^{-1}$ | [14] |
| $\mu_I$ | Additional mortality rate of infected red blood cells | 0.15 | $T^{-1}$ | |
| $\mu_X$ | Mortality rate of merozoites | 0.125 | $T^{-1}$ | [17] |
| $\delta_1$ | Number of merozoites invading a red blood cell | 1 | $[X][H]^{-1}$ | [17] |
| $\gamma_X$ | Rate of antiparasitic action of platelets in merozoites | $*1.0 \times 10^{-9}$ | $[P]^{-1}T^{-1}$ | Assumed |
| $\tilde{\gamma}_X$ | Rate of antiparasitic action of giant platelets in merozoites | $*1.0 \times 10^{-7}$ | $[P]^{-1}T^{-1}$ | Assumed |
| $\gamma_a$ | Rate of antiparasitic action of platelets in infected red blood cells | $*1.0 \times 10^{-9}$ | $[P]^{-1}T^{-1}$ | Assumed |
| $\tilde{\gamma}_a$ | Rate of antiparasitic action of giant platelets in infected red blood cells | $*1.0 \times 10^{-7}$ | $[P]^{-1}T^{-1}$ | Assumed |
| $K_2$ | Production rate of megakaryocytes precursor cells | $*5.0 \times 10^5$ | $[M]^{-1}T^{-1}$ | Assumed |
| $\mu_Z$ | Mortality rate of megakaryocytes precursor cells | $1/20$ | $T^{-1}$ | Assumed |
| $\sigma_1$ | Differentiation rate to megakaryocyte | 0.8 | $T^{-1}$ | Assumed |
| $\sigma_2$ | Differentiation rate to megakaryocyte producing giant platelets | 0.2 | $T^{-1}$ | Assumed |
| $\mu_1$ | Mortality rate of megakaryocyte | 0.1 | $T^{-1}$ | Assumed |
| $\epsilon_1$ | Rate of passage of megakaryocytes into the platelet compartment | 0.8 | $T^{-1}$ | [10] |
| $\epsilon_2$ | Rate of recruitment of megakaryocytes $M_1$ become megakaryocyte $M_2$ | 0.2 | $[P]^{-1}T^{-1}$ | Assumed |
| $\mu_2$ | Mortality rate of megakaryocyte producing giant platelets | $1/12$ | $T^{-1}$ | Assumed |
| $\epsilon_3$ | Rate of passage of megakaryocytes into the giant platelets compartment | 0.9 | $T^{-1}$ | Assumed |
| $\delta_2$ | Number of platelets released by megakaryocyte | 1000 | $[P][M]^{-1}$ | [7] |
| $\mu_{P_1}$ | Mortality rate of platelets | $1/7$ | $T^{-1}$ | [18] |
| $\delta_4$ | Number of platelets that bind to an infected red blood cell | 100 | $[P][H]^{-1}$ | Assumed |
| $\delta_6$ | Number of platelets acting on a merozoite | 10 | $[P][X]^{-1}$ | Assumed |
| $\delta_3$ | Number of giant platelets released for one megakaryocyte $M_2$ | 100 | $[P][M]^{-1}$ | Assumed |
| $\mu_{P_2}$ | Mortality rate of giant platelets | $1/15$ | $T^{-1}$ | Assumed |
| $\delta_5$ | Number of giant platelets that bind to an infected red blood cell | 10 | $[P][H]^{-1}$ | Assumed |
| $\delta_7$ | Number of giant platelets acting on a merozoite | 10 | $[P][X]^{-1}$ | Assumed |

## 3 Analysis

We performed the steady-state analysis in order to evaluate the platelet antiparasitic action during malaria infection. This section is divided into two main parts. In the first part, we study the case where there is no antiparasitic activity of normal or giant platelets. Hence, there are only normal platelets circulating in the bloodstream without interfering with merozoite infection. That is, malaria infection interacts only with erythrocytes. This part is denoted by “Non-interacting platelets and malaria infection”.

In the second part, we analyze both normal and giant platelet antiparasitic activity, how they interact with malaria infection. This part is denoted by “Platelets antiparasitic action “. To investigate the individual contribution of normal platelets and the joint action on the infection control, we analyze the two cases: (i) normal platelet acts against merozoites and infected red blood cells, and (ii) joint action normal and giant platelets.

### 3.1 Non-interacting platelets and malaria infection

Here, platelets do not have antiparasitic action. Therefore, we have two decoupled systems, a system that models the dynamics of normal platelet production and another related to the interaction of merozoites with the target cells.

For the system that models the dynamics of platelet production, we have

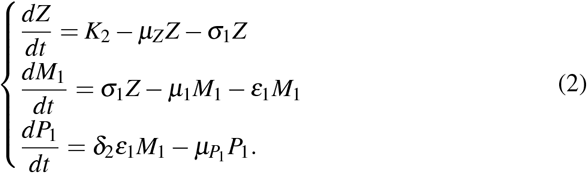

In this case, we have only one equilibrium point given by 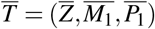, where 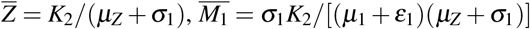, and 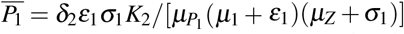.

By the change of variables 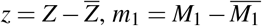, and 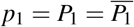, we can prove that *T* is globally asymptotically stable. For more details, see Appendix A.

The system that models the interaction between merozoites and red blood cells without antiparasitic action of platelets is given by

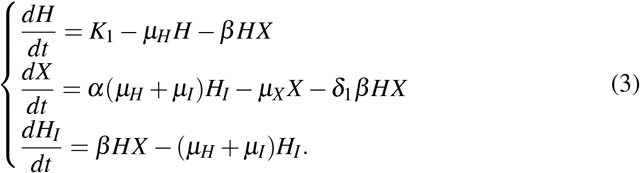

The merozoites-free equilibrium (MFE), 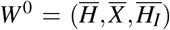, is given by *W* ^0^ = (*K*_1_*/µ*_*H*_, 0, 0). The characteristic polynomial corresponding to the Jacobian matrix evaluated at *W* ^0^ has one negative root −*µ*_*H*_ and two roots given by the solutions of the second-order polynomial

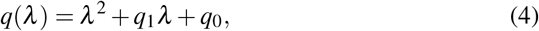

where the coefficients are,

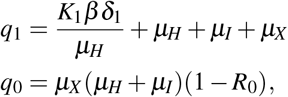

and *R*_0_ is given by,

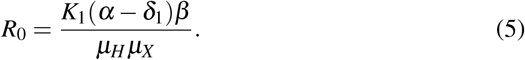

Applying the Routh-Hurwitz criteria [15], MFE is locally asymptotically stable if *R*_0_ *<* 1 and unstable if *R*_0_ > 1. Therefore, the local stability of this point is stated in Theorem 1.

#### Theorem 1

*The merozoites-free equilibrium point W* ^0^ *is locally asymptotically stable if R*_0_ *<* 1, *and unstable if R*_0_ > 1, *where R*_0_ *is given by (5)*.

***Proof:*** *The polynomial q*(*λ*), *which is given by equation (4), satisfies all Routh-Hurwitz conditions if R*_0_ *<* 1, *that is, q*_1_ > 0 *and q*_0_ > 0. □

With respect to *R*_0_, it can be rewritten as 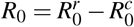, where

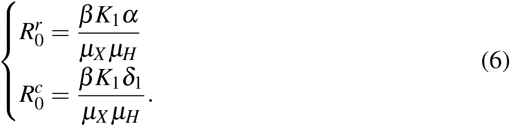

Notice that [(*β K*_1_*/µ*_*H*_)*α/µ*_*X*_] represents the number of new merozoites produced by one merozoite introduced into the bloodstream (containing a susceptible erythrocyte population of size *K*_1_*/µ*_*H*_) free of this parasite. The second term, 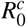, is the number of merozoites consumed by the infected erythrocyte during the cell infection process.

Therefore, *R*_0_ is the net number of merozoites produced by invading merozoites in the early stage of malaria infection.

The merozoites-presence equilibrium (MPE), which is 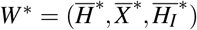, has the coordinates given by

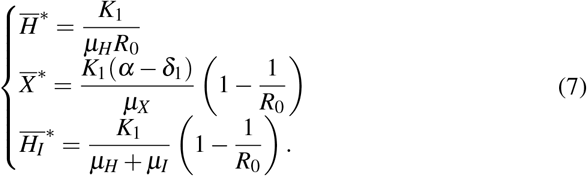

Therefore, MPE exists only if *R*_0_ > 1 due to the biological condition of existence. The characteristic polynomial corresponding to the Jacobian matrix evaluated at *W* ^∗^ is given by

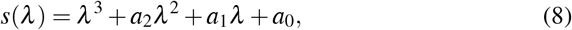

where

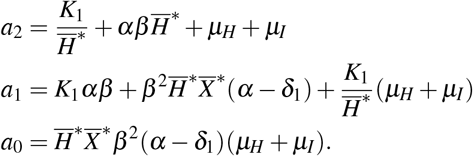

The local stability of *W* ^∗^ is stated in Theorem 2.

#### Theorem 2

*The merozoites-presence equilibrium point W* ^∗^ *is locally asymptotically stable if R*_0_ > 1, *and unstable if R*_0_ *<* 1.

***Proof***. *See Appendix B*.

To obtain global stability of *W* ^0^ and *W* ^∗^, we used the direct method of Lyapunov. The proofs were adapted from Gómez (2016) [4].

#### Theorem 3

*If R*_0_ *<* 1 *then merozoite-free equilibrium W* ^0^ *is globally asymptotically stable*.

***Proof***. *See Appendix C*.

#### Theorem 4

*The merozoite-presence equilibrium W* ^∗^ *is globally asymptotically stable if R*_0_ > 1 *and* 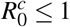.

***Proof***. *See Appendix D*.

We showed that MPE is globally asymptotically stable only if *R*_0_ > 1 and 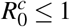. The additional restriction, 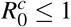, appears because the model considered the cell invasion by the parasite necessary to occur the erythrocyte infection. If we had considered only the contact between the parasite and the erythrocyte to occur the infection, there was no term *δ*_1_*β HX*. Thus, we would have a classic model of Mathematical Epidemiology, and *W* ^∗^ would be globally asymptotically stable, without additional restriction, only *R*_0_ > 1.

### 3.2 Platelets antiparasitic action

Here, we consider the antiparasitic action of both normal and giant platelets. The merozoites-free steady state is given by 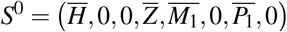, where the nonnull coordinates values are the same as appeared in *W* ^0^ and 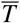.

The characteristic polynomial corresponding to the Jacobian matrix evaluated at *S*^0^ has six negative roots: 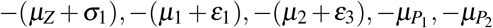 and −*µ*_*H*_; and two roots are given by the solutions of second-order polynomial *w*(*λ*) = *λ*^2^ + *w*_1_*λ* + *w*_0_, where

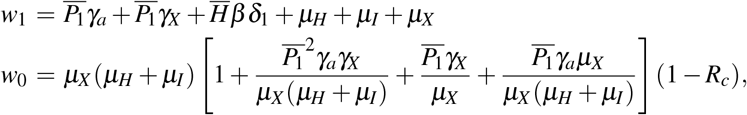

wherein *R*_*c*_ is given by

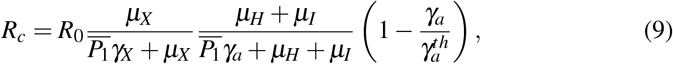

and 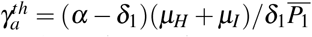.

Applying the Routh-Hurwitz criteria, *S*^0^ is locally asymptotically stable if *R*_*c*_ *<* 1, and unstable if *R*_*c*_ > 1.

#### Theorem 5

*The merozoites-free equilibrium point S*^0^ *is locally asymptotically stable if R*_*c*_ > 1, *and unstable if R*_*c*_ *<* 1.

***Proof***. *The proof is analogue of the proof in Theorem 1. So, as w*_1_ *and w*_0_ *are positive values, the equilibrium point S*^0^ *is locally asymptotically stable if R*_*c*_ *<* 1 *and unstable if R*_*c*_ > 1.□

Let us interpret the parameter *R*_*c*_ given by (9). Considering 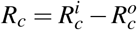, where 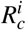 and 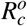 are defined by

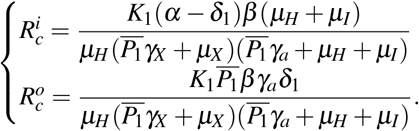

The parameter 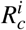 has the following interpretation. Consider that one merozoite during its lifespan 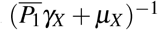, which suffers a decrease due to normal platelets’ action, infects with a rate *β* one susceptible red blood cell in the bloodstream (containing a susceptible erythrocyte population of size *K*_1_*/µ*_*H*_). This infected erythrocyte has a probability 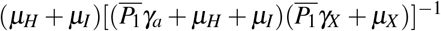 of surviving during its lifespan. If this occurs, this cell will release to the bloodstream *α* new merozoites. To the parameter 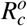, we have the following interpretation, *δ*_1_ merozoites during their lifespan 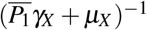 at a rate *β* infect one red blood cell in the bloodstream (containing a susceptible erythrocyte population of size *K*_1_*/µ*_*H*_). This infected erythrocyte has a probability 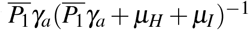 of suffering antiparasitic action.

As a result, the threshold *R*_*c*_ is the net number of merozoites produced by one invading merozoite on the occurrence of antiparasitic action of normal platelets.

Observe that, if 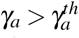, then we have *R*_*c*_ *<* 0. Thus, the action of normal platelets on infected red blood cells is much more efficient than their action on the merozoites, as it is able to make the threshold *R*_*c*_ negative. The negative value of *R*_*c*_ shows that the infection would already be controlled because no new merozoite would be produced from the primary infection. It is also important to emphasize that if all *R*_*c*_ factors are positives, then *R*_*c*_ is less than *R*_0_. It shows that the antiparasitic action of normal platelets decreases the net number of merozoites produced by one invading merozoite.

The merozoites-presence equilibrium *S*^∗^ = (*H*^∗^, *X* ^∗^, *H*_*I*_^∗^, *Z*^∗^, *M*_1_^∗^, *M*_2_^∗^, *P*_1_^∗^, *P*_2_^∗^) has the coordinates given by

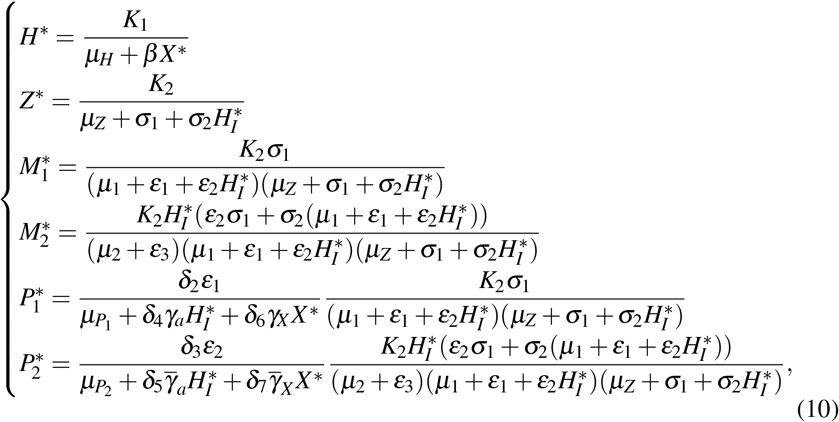

where *X* ^∗^ and 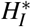 are solution of the system of equations

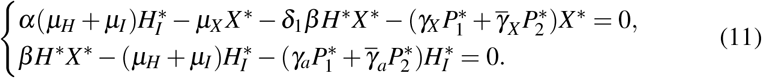

Due to the system (11) be strongly nonlinear, we did not obtain all coordinates of *S*^∗^. So, in the next subsections, we analyze the existence and stability of *S*^∗^ restricted to two different cases: (i) Antiparasitic action of normal platelets without giant platelets and (ii) Joint antiparasitic action of both normal and giant platelets.

#### 3.2.1 Antiparasitic action of normal platelets without giant platelets

In this case, only normal platelets have antiparasitic action, and the giant platelets are not being produced. Simplifying the system of equations (11), we obtain *S*^∗^ = (*H*^∗^, *X* ^∗^, *H*_*I*_^∗^, *Z*^∗^, *M*_1_^∗^, 0, *P*_1_^∗^, 0), wherein *H*^∗^, *Z*^∗^, *M*_1_^∗^, and *P*_1_^∗^ are given by (10) and *H*_*I*_^∗^ is given by

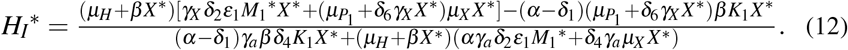

Substituting all equilibrium values of (10) and (12) in the first equation of (11), we obtain an equation with only the variable *X* ^∗^. Simplifying this equation, we obtain *p*(*X* ^∗^) = *f* (*X* ^∗^) −*g*(*X* ^∗^) = 0, where *f* (*X* ^∗^) and *g*(*X* ^∗^) are fourth-degree polynomials. The expressions are presented in Appendix E. By the Descartes’ rule of sign, we found that if 0 *< R*_*c*_ *<* 1, then there are zero or two positive roots (biologically viable) for *X* ^∗^. If *R*_*c*_ > 1 there is a unique positive root (biologically viable) for *X* ^∗^.

We analyze two cases to evaluate the dependence of the parameters *γ*_*a*_ and *γ*_*X*_ on the existence of positive equilibrium points for *X* ^∗^ with *R*_*c*_ ∈ [0, 1]. Firstly, we analyze the influence of normal platelets being added to infected red blood cells, and after that, the influence of normal platelets being added into merozoites. The analysis of both cases will be through bifurcation diagrams, where we fixed all parameters (Table 2), except *β, γ*_*a*_, and *γ*_*X*_, in order to explore the influence of *γ*_*a*_ and *γ*_*X*_ for different values of the infection parameter *β*.

Following the same idea, we analyze the influence of *γ*_*X*_ in the merozoites-presence equilibrium point. We found a similar behavior to the diagrams varying *γ*_*a*_ so that the diagrams will not be shown here.

Figure 1 shows that increasing the value of *β* increases the concentration of merozoite, which was expected. In addition, the higher *β*, the higher the value of *γ*_*a*_ needed to make the value of *R*_*c*_ smaller than one, that is, eliminating the parasite concentration. Here, we found a forward bifurcation, that is, when *R*_*c*_ *<* 1 there only is MFE point *S*^0^, which is LAS and if *R*_*c*_ > 1 a LAS MPE point *S*^∗^ appears and the MFE point *S*^0^ becomes unstable.

**Fig. 1:**
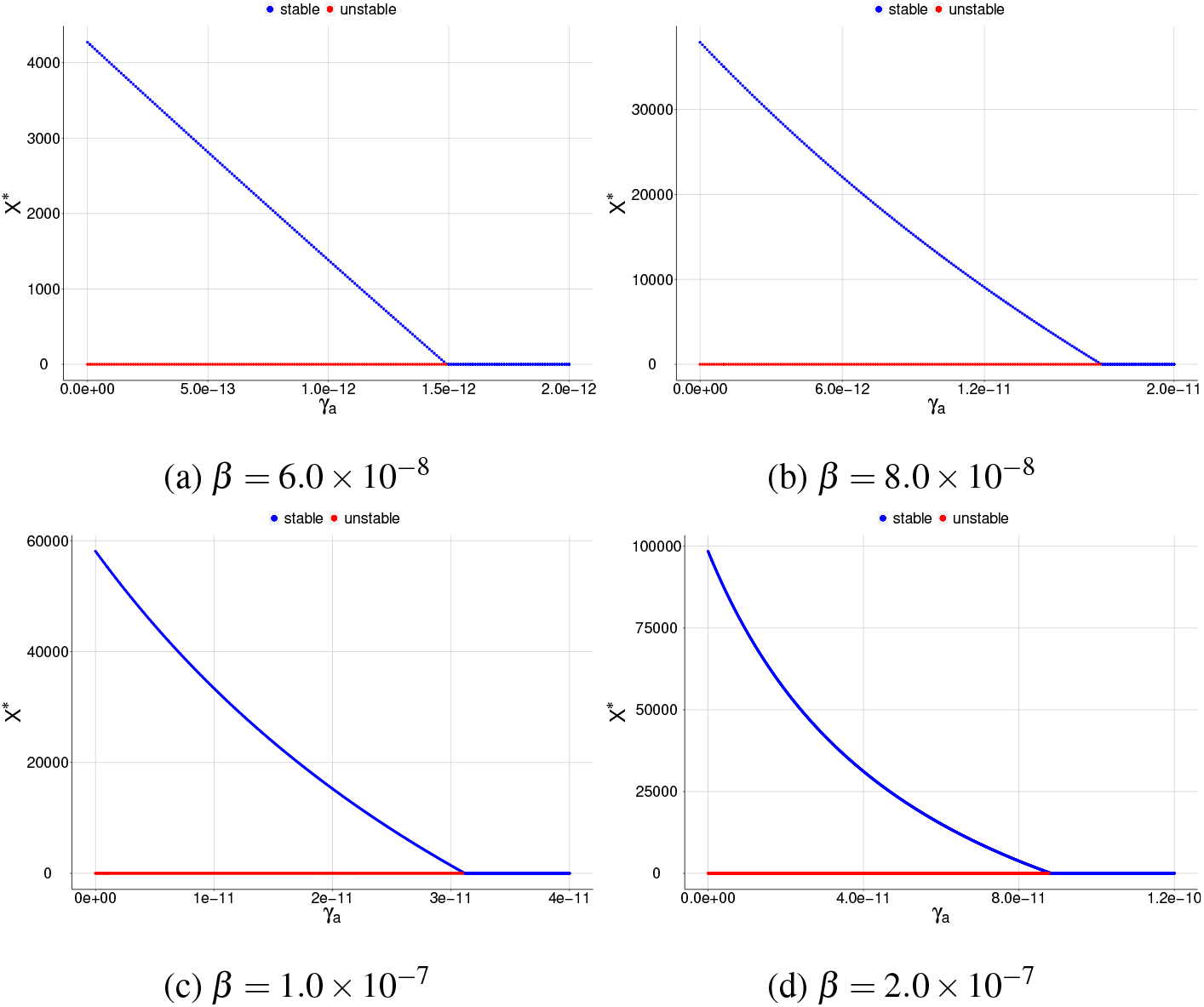
Bifurcation diagrams for *X* ^∗^, where *γ*_*a*_ is chosen as a bifurcation parameter. All plots shows a reverse forward bifurcation. For (a), (b), (c), and (d), the blue color corresponds to the stable equilibrium and the red color corresponds to the unstable equilibrium.

Appendix F illustrates a case with low platelet production. In that case, a backward bifurcation could appear. It is possible to have two MPE points when *R*_*c*_ is less than one. Also, depending on the *β* and *γ* values, oscillatory behaviors may arise.

#### 3.2.2 Joint antiparasitic action of both normal and giant platelets

Here, we analyze the joint action of normal and giant platelets on combating merozoites infection. To evaluate the antiparasitic action of giant platelets, we take four cases for the action of giant platelets on infected red blood cells and merozoites so that the values of 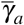 and 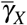 increase: (i) 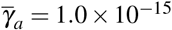 and 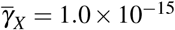, (ii 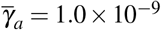 and 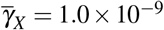, (iii) 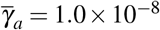 and 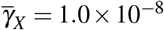, and (iv) 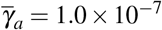 and 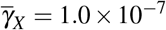.

In order to investigate the effect of giant platelets to assist antiparasitic action of normal platelets, we consider the case of low production rate for megakaryocyte precursor cells; that is, we have a low platelet production. This case is presented in Appendix F.

The first case that will be portrayed is when only normal platelets are acting against merozoite infection. We chose a case from item (c) of Figure 3, with *γ*_*a*_ = 7.0 × 10^−9^, which has only unstable equilibrium points.

**Fig. 2:**
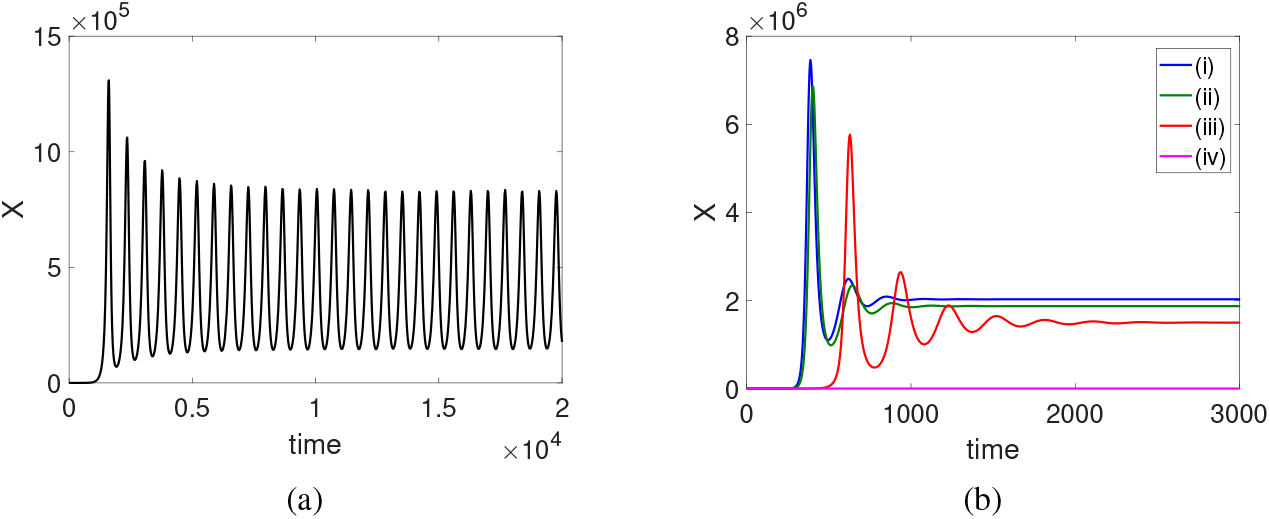
Evaluation of the effect of giant platelet action on oscillatory behavior of merozoite concentration. (a) It shows a oscillatory behavior for *X* ^∗^, when there is an antiparasitic action of normal platelets. (b) Inclusion of different values ((i) 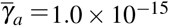 and 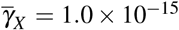, (ii) 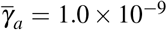 and 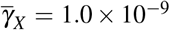, (iii) 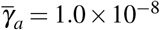 and 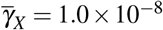, and (iv) 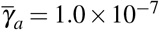 and 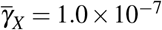) for the antiparasitic action of giant platelets in the behavior shown in item (a). As initial condition, we considered the coordinates of the merozoites-free equilibrium point, except for merozoites, which was considered *X* (0) = 1.

**Fig. 3:**
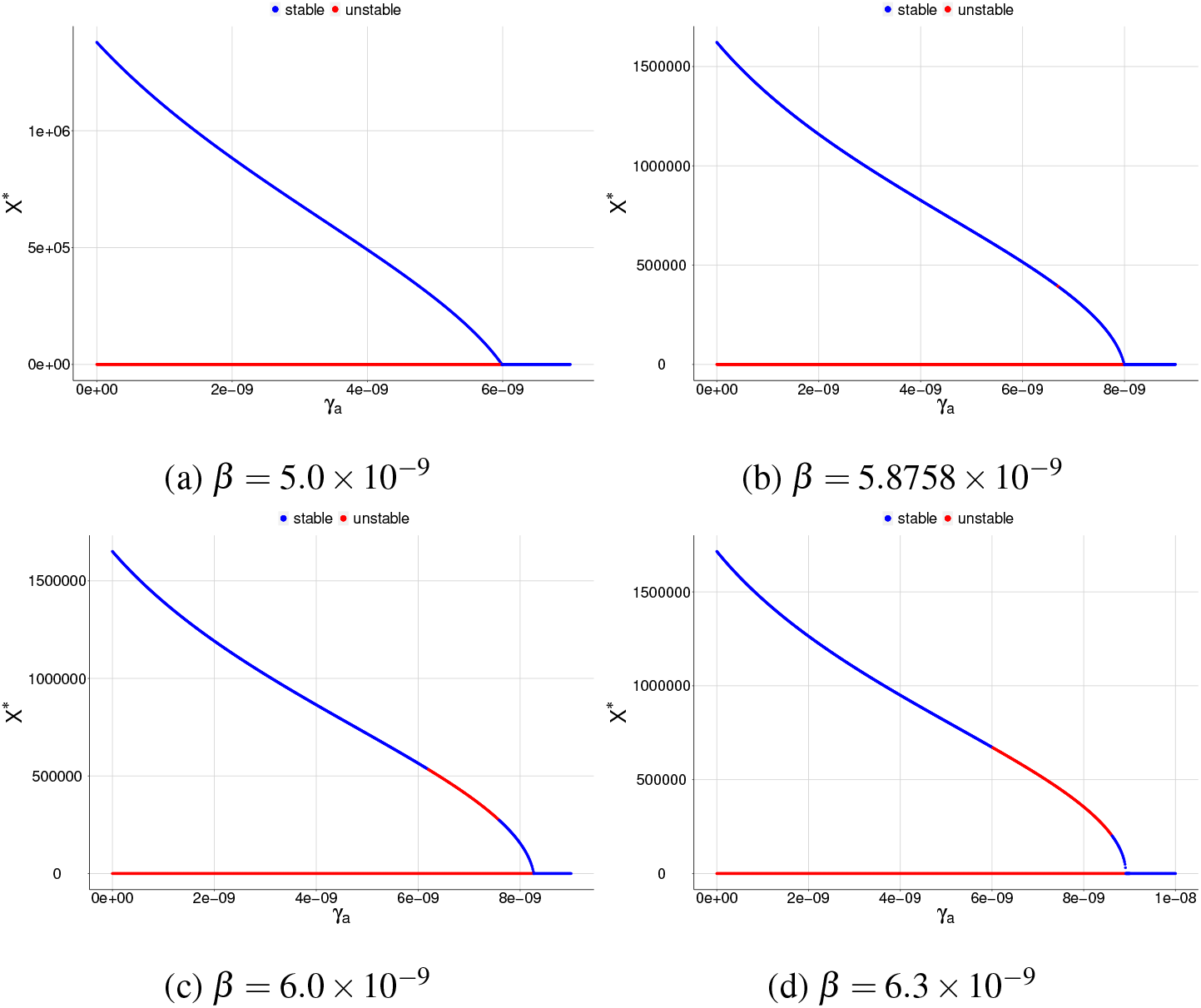
Bifurcation diagrams for *X* ^∗^, where *γ*_*a*_ is chosen as a bifurcation parameter in the case of low platelet production. All graphs show a reverse forward bifurcation. For (a), (b), (c), and (d), the blue color corresponds to the stable equilibrium, and the red color corresponds to the unstable equilibrium.

In Figure 2b, for a small value (blue color), the giant platelet action contributes to eliminating the oscillatory behavior of the dynamics of the system, stabilizing their values. As the antiparasitic action of giant platelets on infected red cells and merozoites increases, parasite concentration decreases, so in item (d), we have a convergence close to zero for *X*.

Thus, even in the case of low production of megakaryocyte precursor cells, with the antiparasitic action of giant platelets, it is possible to decrease the parasite concentration or even control it.

Therefore, giant platelets assisting the existing normal platelet antiparasitic action assist malaria infection control.

## 4 Discussion

We proposed a mathematical model to evaluate the antiparasitic action of both normal and giant platelets against merozoites and infected red blood cells during malaria infection. Our model showed that merozoites’ concentration when they suffered antiparasitic action was lower than the case without the antiparasitic response of platelets.

Without normal and giant platelet antiparasitic action, the model had two equilibrium points, the merozoite-free equilibrium *S*^0^, which was globally asymptotically stable if the threshold *R*_0_ is less than one, and unstable if *R*_0_ is greater than one. The merozoites-presence equilibrium *S*^∗^ was locally asymptotically stable if *R*_0_ > 1 and unstable if *R*_0_ *<* 1. The threshold *R*_0_ represents the net amount of new merozoites that one merozoite can produce. Therefore, if *R*_0_ *<* 1 even in a situation of colossal parasite inoculation, the host could fight against the merozoite and eliminate the infection. However, if *R*_0_ > 1, the introduction of only a single merozoite led to the persistence of parasitic concentration in the host.

When we included to the model the antiparasitic action of platelets, we obtained a new threshold, *R*_*c*_, which was related to *R*_0_, with *R*_*c*_ ≤ *R*_0_. The threshold *R*_*c*_ represents the net number of new merozoites that one merozoite can produce under the effect of normal platelet antiparasitic action. Also, the antiparasitic action of normal platelets contributed to decreasing *R*_0_. Thus, in a case with *R*_0_ > 1, we can have a *R*_*c*_ *<* 1, leading to population dynamics converge to *S*^0^, but without antiparasitic action of platelets would converge to *S*^∗^. Moreover, in the threshold *R*_*c*_, there were no giant platelet antiparasitic action parameters. Our model considered that the giant platelets were produced in the bone marrow only after the red blood cell infection by merozoites. In the existence of infected red blood cells, the megakaryocyte also produces this platelet type. Therefore, the antiparasitic action of normal platelets occurred as a primary response against the infection. The antiparasitic action of giant platelets occurred as a secondary response to assist the normal platelet antiparasitic action.

The antiparasitic action of normal platelets on infected erythrocytes, by means of the parameter *γ*_*a*_ proved to be able to control infection by itself, as long as *γ*_*a*_ is greater than 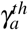 the threshold *R*_*c*_ will be negative. Hence, no infection would persist even if we introduce a huge merozoite concentration in the bloodstream. However, this effect was not possible using only the antiparasitic action of normal platelets on merozoites, utilizing the parameter *γ*_*X*_.

Considering a low platelet production, which may lead to a case of thrombocytopenia, a change of a forward to backward bifurcation as we increase the value of *β* occurred. In this case, we obtained the possibility of having up to three equilibrium points, *S*^0^, and two positive equilibrium points.

Depending on the parameters *γ*_*a*_ and *γ*_*X*_, there was a possibility of appearing a limit cycle, which led to an oscillatory behavior for the population dynamics. The introduction of the giant platelet action to the case of a limit cycle could eliminate this oscillatory behavior. The higher the giant platelet’s action, the lower the concentration of the parasite.

When we considered a normal platelet production, the system (1) had only two equilibrium points. In that case, we had a classical behavior for the existence and stability of the equilibrium points; that is, we obtained only one positive equilibrium point, which was locally asymptotically stable if *R*_*c*_ is higher than one and unstable if *R*_*c*_ is less than one.

## 5 Conclusion

In conclusion, through a simple mathematical model, we determined the antiparasitic effect that normal and giant platelets can have on infected red blood cells and circulating merozoites. We emphasize that our goal with this model was not to have a quantitative framework but to present a theoretical approach to the dynamic behavior between antiparasitic action of platelets and the course of malaria infection to elucidate the role of platelets during malaria infection. In the absence of platelet antiparasitic activity, high levels of parasitic concentration were achieved, as expected. The antiparasitic action of normal platelets contributed to decreased parasite concentration levels in the bloodstream or even eliminated the parasite. However, the low production of platelets led to the occurrence of backward bifurcation in this model. It implied the existence of a merozoite-presence equilibrium point even when the threshold parameter for the net number of merozoites produced was below the unit, which showed that the malaria infection could persist or be controlled in a case of low platelet count. Also, in the case of low production of platelet, a limit cycle may appear. The appearance of giant platelets occurred to assist the antiparasitic action of normal platelets to eliminate the limit cycle and contributed to the control of the parasite. Models that take into account the immune system and antimalarial drugs merit further investigations.

## Data Availability

The data used are available online.

https://www.who.int/en/news-room/fact-sheets/detail/malaria

## Acknowledgements

The authors are grateful to Coordination of Superior Level Staff Improvement (CAPES) - Finance Code 001 by the financial support.

## Conflict of interest

The authors declare that they have no conflict of interest.

## A Global stability of the equilibrium point *T̅*

Using the change of variables 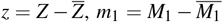 and 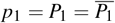, the system (2) results in a homogeneous linear system. The solution is given by

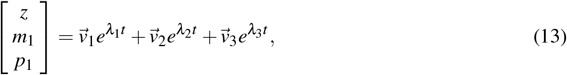

where 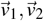, and 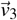 are eigenvectors associated with the eigenvalues *λ*_1_ = −(*µ*_*Z*_ + *σ*_1_), *λ*_2_ = −(*µ*_1_ + *ε*_1_), and *λ*_3_ = −*µ*_*P*_1, respectively.

Notice that when *t* → ∞, the solution (13) tends to the null vector, so we concluded that 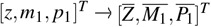. Therefore, the system converges to 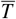 for any initial condition, resulting in a globally asymptotically stability.

### B Local stability of the point *W* ^∗^

To show the local stability of the point *W* ^∗^, we proved that the coefficients of the characteristic polynomial *s*(*λ*), given by equation (8), satisfy the Routh-Hurwitz conditions: *a*_2_ > 0, *a*_1_ > 0, *a*_0_ > 0 and *a*_2_*a*_1_ > *a*_0_. It’s easy to verify that *a*_2_, *a*_1_, and *a*_0_ are positives when *R*_0_ > 1. To prove the condition *a*_2_*a*_1_ > *a*_0_, we consider *a*_2_*a*_1_ − *a*_0_, and this term proved to be positive.

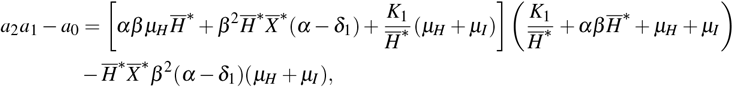

rewriting, we found that

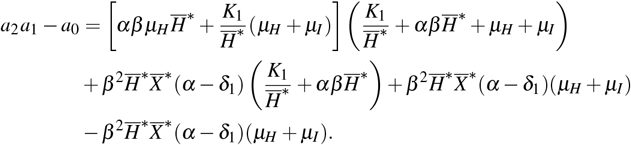

The two last terms are canceled, so there are only positive terms. Thus, we concluded that *a*_2_*a*_1_ > *a*_0_.

### C Proof of global stability of *W* ^0^

Consider *f* a function defined by *f* : *Ω*_1_ → ℝ, such that

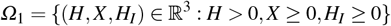

and

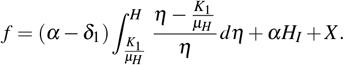

As *W* ^0^ = (*k*_1_*/µ*_*H*_, 0, 0), observe that *f* (*W* ^0^) = 0 and *f* > 0 in *Ω*_1_ − {*W* ^0^}. In addition,

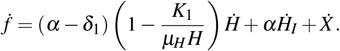

Replacing the expressions for 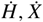 and 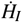 we find that

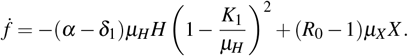

Therefore, 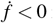 in *Ω*_1_ −{*W* ^0^} if and only if *R*_0_ ≤ 1. It is concluded that merozoites-free equilibrium point *W* ^0^ is globally asymptotically stable.

### D Proof of global stability of *W* ^∗^

Consider the function *g* defined by *g* :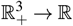, where

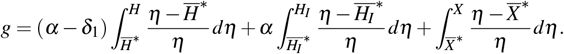

Therefore, the derivative function is given by

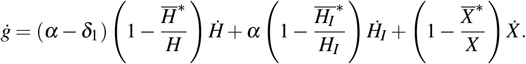

Simplifying the terms of the above equation, we have

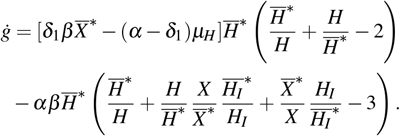

Being the arithmetic mean greater than or equal to the geometric mean, that is,

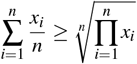

we have 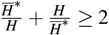 and 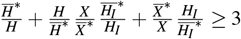.

Thus, if 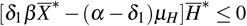 we have *ġ <* 0 because

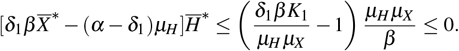

Hence, we have *ġ* < 0 in 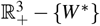. In addition, *g*(*W* ^∗^) = 0 and *g* > 0 in 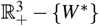. Therefore, the merozoites-presence equilibrium, *W* ^∗^, is globally asymptotically stable.

### E Polynomials *f* and *g* shown in section 3.2.1

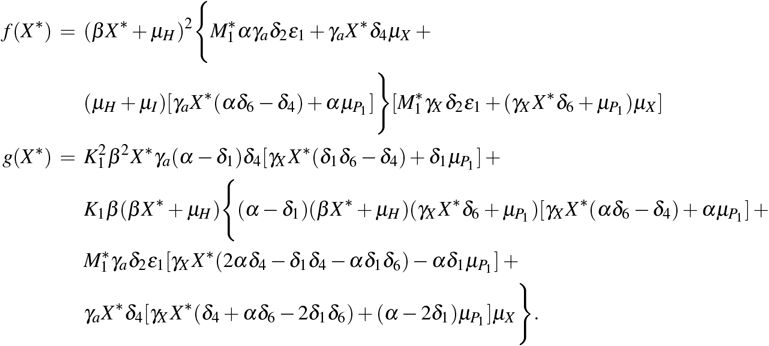

### F A case of normal platelet antiparasitic action with a low production rate for megakaryocyte precursor cells

Here, the megakaryocyte precursor cells have a low production rate; we consider *K*_2_ = 3.0 × 10^3^. Through bifurcation diagrams varying the parameters *γ*_*a*_ and *γ*_*X*_ for different values of *β*, we analyze the antiparasitic action of normal platelets considering that there is a low platelet production.

we obtain that the graphs only have a reverse forward bifurcation for low *β* values, and the merozoites-presence equilibrium point is stable for 0 *< R*_*c*_ *<* 1.

In Figure 3, we obtain that the graphs only have a reverse forward bifurcation for low *β* values, and the merozoites-presence equilibrium point is stable for 0 *< R*_*c*_ *<* 1. However, as we increase *β* and high *γ*_*a*_ values an unstable region appears to the merozoites-presence equilibrium point. As we increase the value of *β*, the unstable region grows. Also, two positive equilibrium points may appear. In this case, the bifurcation diagrams have reverse backward bifurcation, which is given by Figures 4 to 5.

**Fig. 4:**
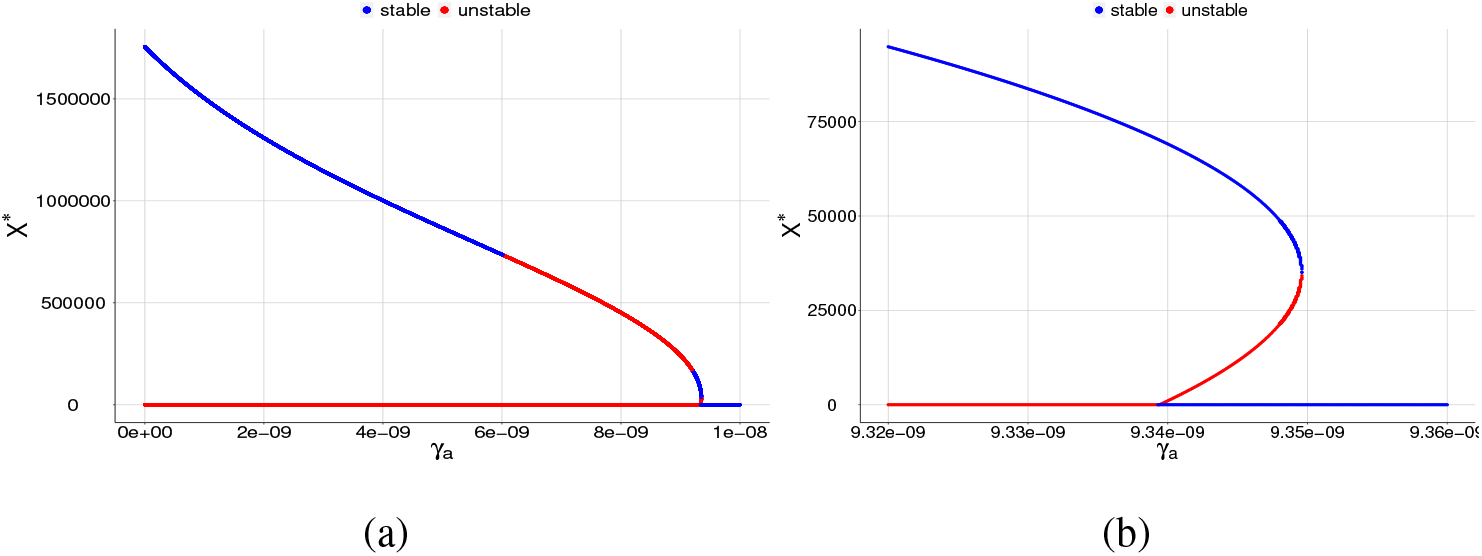
Bifurcation diagrams for *X* ^∗^, where *γ*_*a*_ is chosen as a bifurcation parameter and *β* = 6.5 × 10^−9^. The blue color corresponds to the stable equilibrium and the red color corresponds to the unstable equilibrium point. For (a) the graph shows a reverse backward bifurcation diagram and (b) same graph as item (a) but restricted to a smaller interval for the parameter *γ*_*a*_ in order to see the two positive equilibrium points.

**Fig. 5:**
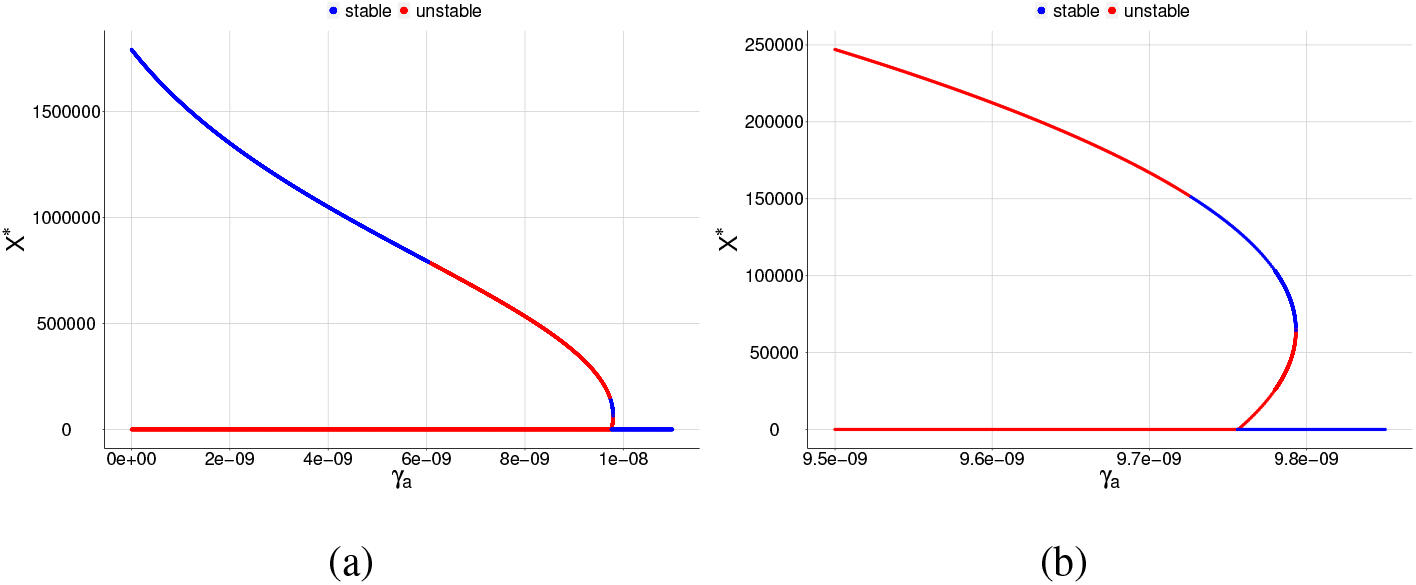
Bifurcation diagrams for *X* ^∗^, where *γ*_*a*_ is chosen as a bifurcation parameter and *β* = 6.7 × 10^−9^. The blue color corresponds to the stable equilibrium and the red color corresponds to the unstable equilibrium point. For (a) the graph shows a reverse backward bifurcation diagram and (b) same graph as item (a) but restricted to a smaller interval for the parameter *γ*_*a*_ in order to see the two positive equilibrium points.

From Figures 3 and 4, we can see that as we increase the value of *β*, we pass from the existence of two equilibrium points to the presence of three equilibrium points. Depending on the value of *γ*_*a*_, it is possible to have the following situations:

- there are two equilibrium points: a stable (non-zero equilibrium point) and the other is unstable (merozoites-free equilibrium point),
- there are only two unstable equilibrium points, according to the item (d) of Figure 3 (merozoites-free equilibrium point and a non-zero equilibrium point),
- there are three equilibrium points, one stable and two unstable, according to the item (b) of Figure 7 or two stable and one unstable equilibrium points, according to the item (b) of Figure 4.

In the case of three equilibrium point existence, we found that the stable region decreases when we increase *β*. Lastly, we can see that only to low values of *γ*_*a*_, the merozoites-presence equilibrium point is stable; otherwise, it is unstable.

**Fig. 6:**
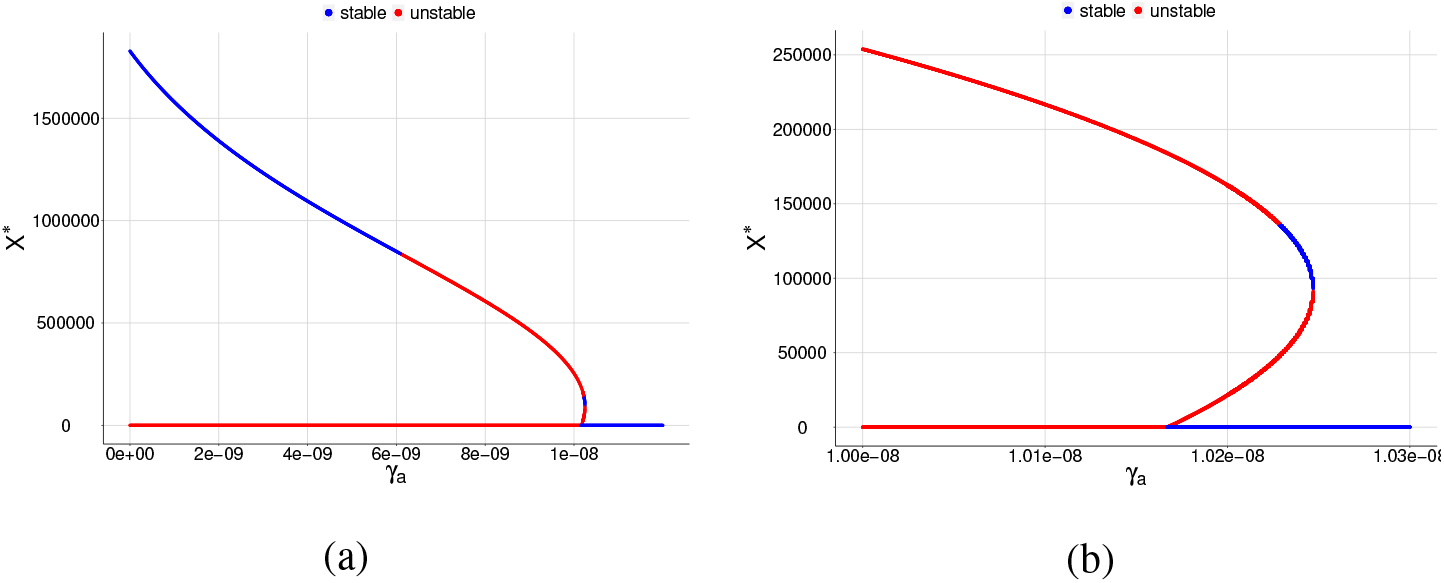
Bifurcation diagrams for *X* ^∗^, where *γ*_*a*_ is chosen as a bifurcation parameter and *β* = 6.9 × 10^−9^. The blue color corresponds to the stable equilibrium and the red color corresponds to the unstable equilibrium point. For (a) the graph shows a reverse backward bifurcation diagram and (b) same graph as item (a) but restricted to a smaller interval for the parameter *γ*_*a*_ in order to see the two positive equilibrium points.

**Fig. 7:**
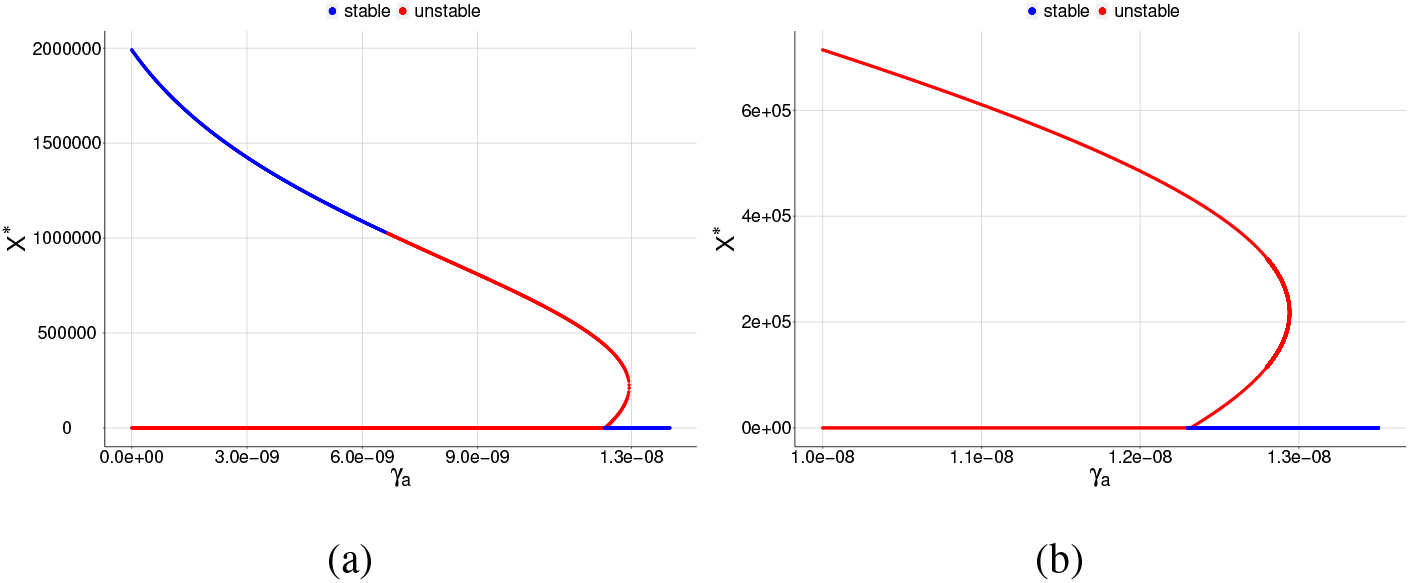
Bifurcation diagrams for *X* ^∗^, where *γ*_*a*_ is chosen as a bifurcation parameter and *β* = 8.0 × 10^−9^. The blue color corresponds to the stable equilibrium and the red color corresponds to the unstable equilibrium point. For (a) the graph shows a reverse backward bifurcation diagram and (b) same graph as item (a) but restricted to a smaller interval for the parameter *γ*_*a*_ in order to see the two positive equilibrium points.

When *γ*_*a*_ surpasses a critical value 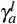, the merozoites-presence equilibrium point becomes unstable, and a cycle limit appears, which can be numerically assessed through the eigenvalues of the associated Jacobian matrix. We found a pair of complex eigenvalue with zero real part. For another value of *γ*_*a*_, in this case, we denote by 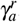, we found a limit cycle, and when *γ*_*a*_ surpasses 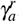, the merozoites-presence equilibrium point becomes stable again.

As presented, a Hopf bifurcation appears in the diagrams since it presents a complex eigenvalue pair whose real part is null. To illustrate this situation, we chose item (c) of Figure 3. Figure 8 illustrates the value of (a) the real part and (b) imaginary part, which are assumed in the complex eigenvalue pair as we increase the value of *γ*_*a*_.

**Fig. 8:**
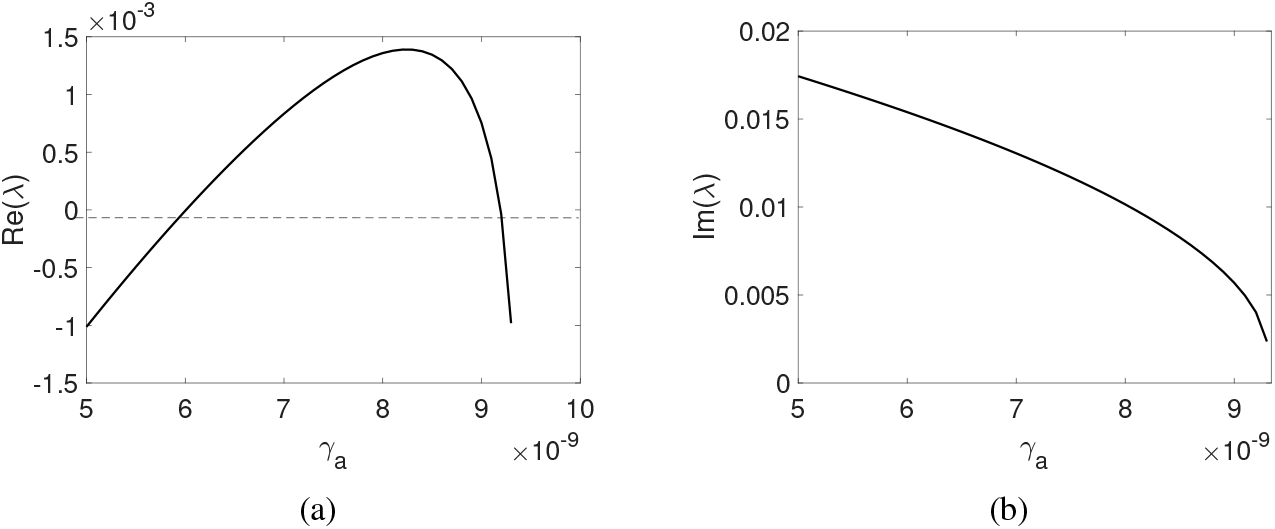
An example of Hopf bifurcation that appears in the model. (a) Real part and (b) imaginary part of the complex eigenvalue pair varying the parameter *γ*_*a*_, respectively.

We found an eigenvalue with null real part for 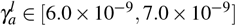 and after for 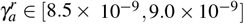, showing Hopf bifurcations when varying *γ*_*a*_. In the other bifurcation diagrams, it is also possible to find two Hopf bifurcations; they arise precisely at the point where the change of stability occurs.

## Notes

### Competing Interest Statement

The authors have declared no competing interest.

### Funding Statement

FAR thanks the Coordination of Superior Level Staff Improvement (CAPES) for the financial support (Finance Code 001).

### Author Declarations

This paper does not include animal and/or human trials

